# Dissecting age-stratified immunity to different dengue virus serotypes and Zika viruses among children in a highly endemic region in Sri Lanka

**DOI:** 10.1101/2025.06.25.25330279

**Authors:** Shyrar Tanussiya Ramu, Madushika Dissanayake, Chamini Kanatiwela-de Silva, Naduni Dasanthi, Amaya Gunaratne, Saubhagya Danasekara, Laksiri Gomes, Chandima Jeewandara, Nicole L. Achee, John P. Grieco, H. Asitha de Silva, D. S Anoja F. Dheerasinghe, Thomas W. Scott, Amy C. Morrison, Hasitha Aravinda Tissera, Gathsaurie Neelika Malavige

## Abstract

**Background:** Determining serotype-specific age stratified dengue (DENV) and Zika (ZIKV) seroprevalence rates is crucial for implementing vaccines and vector control strategies. Therefore, we sought to assess age-stratified seroprevalence of monotypic or multi-typic exposure in a community cohort in Sri Lanka.

**Methods:** The DENV-specific serostatus was assessed in 4,161 children aged 4 to 16 years, using an in-house DENV specific IgG ELISA and compared the results with a widely used commercial assay (dengue indirect Panbio IgG ELISA). We also used a multiplexed, microsphere-based serological assay, to characterise monotypic vs multi-typic infections and to differentiate exposure rates to different DENV serotypes and ZIKV in a sub-cohort of children (n=604).

**Results:** By IgG-ELISA DENV seropositivity was 72.34% (n=4,161), and the seroprevalence rate significantly increased with age (Spearman’s r=1.0, p=0.003). The estimated FOI was 0.16 (95% credible interval 0.14–0.17). Of the 604 individuals tested by Luminex, 258 (42.7%) had a monotypic dengue infection, whereas 209 (34.9%) had a multi-typic response. 100 (16.5%) had evidence of a past infection to zika, while 20 (3.33%) children, only had antibodies to the ZIKV. Of the 258 individuals with evidence of a monotypic infection of DENV, DENV2 (56.83%) and DENV1 (30.57%) accounted for most infections. An inverse correlation between exposure to ZIKV and age (Spearmans’s r=-0.72, 0=0.007).

**Conclusions:** 72.3% of children were seropositive for dengue with 42.7% been infected with only one DENV in the past. The data suggests that prior immunity to DENV may reduce the risk of ZIKV infection, which should be further assessed.

## Background

Dengue is the most rapidly emerging mosquito borne viral infection, with 13 million cases reported in 2024 [1]. The burden due to dengue is predicted to further rise due to climate change, rapid urbanization and population displacements [2] and has also been named in the WHO R&D blue print as a pathogen of pandemic potential [3]. Although it has caused outbreaks in many tropical and subtropical countries for over five decades, there are very few innovations been developed in vector control strategies, diagnostics, treatments and in development of vaccines. Although two dengue vaccines have been licenced for prevention of dengue, CYD-TDV was found to have limited efficacy against some DENV serotypes with a potential to cause an increase in hospitalization and severe dengue in dengue naïve individuals [4]. The most recently licenced vaccine, TAK-003 (Qdenga), again is not equally effective against all dengue virus (DENV) serotypes [4]. Further, the WHO has recently recommended that the TAK-003 to be used in children aged 6 to 16 years of age, in high transmission settings, while not recommending the vaccine to be used in low or moderate dengue transmission settings [5]. Therefore, in order to implement vaccine strategies and integrated vaccination-vector control strategies [6], it is crucial to understand the dynamics of dengue transmission in different settings in endemic countries.

Sri Lanka has been experiencing regular dengue outbreaks since 1989 with a gradual rise in the number of cases reported in recent years [7]. Although dengue outbreaks are reported from all over the country, 50% of the cases are reported from the Western province, in the Colombo and Gampaha districts [7]. Based on population studies in 2003, 2013, 2017, 2022, and 2024 conducted in the Colombo district (citation), age seroprevalence curves have been gradually showing a shift to higher seroprevalence rates in younger age groups consistent with a rise in dengue transmission [7, 8]. However, our recent island wide age-stratified dengue seroprevalence study demonstrated significant heterogeneity across different districts in Sri Lanka, with those differences between urban, rural and estate areas [9]. For instance, while the estimated force of infection (FOI) for city of Colombo is 0.149, the FOI for sub-urban areas in Colombo was estimated to be 0.068 [10]. For some districts in Sri Lanka the FOI has estimated to be as low as 0.011 (Badulla, which is predominantly a estate region) and in districts such as Kurunegala (0.012), Matara (0.014) and Kandy (0.013), which are predominantly rural areas [10].

Although, age stratified seroprevalence studies give some indication of the burden of infection in a community, the assays used in these studies (DENV-specific IgG detection assays), detect cross reactive antibodies for other flaviviruses and also cannot differentiate between a past infection due to one DENV serotype (monotypic infections) and infection due to several DENV serotypes (multi-typic infections) [8, 11, 12]. Differentiating the monotypic vs multi-typic exposure to the DENV would provide better granularity in understanding the transmission dynamics of dengue infections. Furthermore, differentiating cross reactive flavivirus antibody responses are important to understand how population immunity to different flaviviruses affect the emergence and spread of dengue outbreaks. For instance, it has been shown that infection with the zika virus (ZIKV) enhanced dengue disease severity during subsequent dengue infections [13]. As IgG antibodies to DENV and ZIKV highly cross react with each other [14], conventional assays that measure either DENV or ZIKV specific IgG would not provide reliable information regarding past infection with either different DENV serotypes of ZIKV.

To understand the dengue transmission dynamics, the rates monotypic or multi-typic DENV infection as well as past infection with ZIKV in Sri Lanka, we studied age stratified seroprevalence in a large cohort of children and then proceeded to use a multiplexed, microsphere-based serological assay (Luminex assay), to differentiate exposure rates to different DENV serotypes and ZIKV in an endemic region in Sri Lanka.

## Materials and Methods

### Study participants

This study was carried out under the Advancing Evidence for the Global Implementation of Spatial Repellents (AEGIS) study, which aims to generate evidence for implementation of novel vector control strategies.

Children (n=4,161), 4 to 16 years of age, were recruited from three divisions (MOH areas) from the Gampaha district, Sri Lanka, which reported a high incidence of dengue. The Medical Officer of Health (MOH) area refers to a specific administrative division, where each area is overseen by a Medical Officer of Health, who is responsible for implementing and managing public health services. This study was conducted as part baseline studies for a vector intervention trial with randomized control trial designed with 30 clusters (10 from each MOH area).

### Sampling technique

All houses within the selected clusters that gave their consent were mapped, and houses with children aged 4-16 years were offered enrollment. Enrollment continued in each cluster until the target sample size of n=130 per cluster was achieved. Blood samples were collected from these children by trained healthcare professionals using standard venipuncture procedures [6]. The collection was carried out in a controlled and sterile environment to ensure the safety and comfort of the children.

### Ethical approval

Ethical approval for the study was obtained from the Ethics Review Committee, University of Kelaniya, Sri Lanka, and also approved by the Institutional Review Boards of the WHO and the University of Norte Dame. All participants were recruited into the study after the provision of informed consent from their parents or guardians, with assent obtained from children where appropriate.

### Development and optimization of an in-house dengue IgG ELISA to determine serostatus of individuals

An in-house ELISA was developed and optimized to measure the presence of DENV-specific IgG antibodies in the children. The in-house ELISA was validated using the Foci reduction neutralization assays (FRNTs)[15] as the gold standard and further compared with a commercial assay (Panbio Dengue Indirect IgG ELISA, Brisbane, Australia), which has been widely used to detect the presence of serostatus of individuals in community sero-surveillance studies in Sri Lanka and elsewhere. Details of the development and validation of the in-house DENV-specific IgG assay are given in supplementary methods.

### The multiplexed, microsphere-based serological assay to determine past infection history

A multiplexed, microsphere-based serological assay (Luminex assay) was carried out as previously described [16] to determine if DENV-seropositive children had been exposed to only one infection (monotypic), many infections (multi-typic) or if they had been exposed to zika. This assay uses antigens of the most variable region of the DENV and ZIKV (EDIII) to differentiate the antibody responses to different DENV serotypes and ZIKV [16]. These assays were developed by the laboratory of Professor Lakshmanane Premkumar from Department of Microbiology and Immunology, University of North Carolina School of Medicine, Chapel Hill, NC, USA and obtained from his laboratory for this study. These assays were carried out on 20% of the samples (n=604), which gave a positive response with the in-house DENV-specific IgG assay. We randomly selected children using stratification by age group and gender to ensure representation across these categories.

Briefly, DENV1-4 and Zika EDIII antigens were first coupled to MagPlex Avidin microspheres (Luminex). The coupled beads were washed and then pooled together into one well of a 96 well plate at the same concentration, after which they were incubated with the selected serum samples to allow binding. The wells were then washed and anti-human total IgG conjugated with Fc-phycoerythrin (Southern Biotech, Birmingham, AL, USA) was added to each well in order to detect the antibodies bound to the beads. After incubation, the wells were washed, and the plate was loaded into a Luminex MAGPIX instrument (xPONENT 4.2 System, USA) to measure the relative antibody (IgG) levels for each antigen (expressed in median fluorescent intensity (MFI) values). A cut-off was set using responses of known negative samples and this cut-off was used to determine positive responses to the antigens. If a positive response was seen only in one DENV antigen, then the result was categorized as a monotypic antibody profile, whereas, if the responses were positive for 2 or more DENV antigens, then the signals for the antigens were normalized and compared at percentages, with the highest signal set to 100%. The positivity threshold for the normalized data was 33% [17] Therefore, if more than one antigen have a signal greater than 33% then it was categorized as a multitypic antibody profile. Those who had responses to the Zika antigen above the cut-off and the threshold were categorized as being positive for Zika.

### Statistical analysis

Statistical analysis was carried out using GraphPad Prism version 10.1. Non-parametric tests were carried out, since the data was not normally distributed. Spearman rank order correlation coefficient was used to determine the correlation between age and seropositivity rates, age and IgG responses and the gender and IgG responses. Spearman’s correlation was also carried out to compare age-stratified seropositivity rates between females and males. Agreement between the in-house and Panbio assays was assessed using Cohen’s kappa coefficient. Additionally, McNemar’s test used to reveal the statistically significant difference between the two assays in identifying seropositive individuals. Age-stratified seropositivity rates were further examined using simple linear regression, with seropositivity rates as the dependent variable and age as the predictor. To evaluate whether the relationship between age and seropositivity rates differed by assay type, an interaction model was fitted. Annual seroconversion rates were estimated by calculating the slope of the best-fit line from the linear regression model across age groups. To estimate the force of infection (FOI), we applied a Bayesian catalytic model assuming a constant FOI across age. The model estimates the cumulative probability of seroconversion as a function of age. The standard form of the catalytic model P(a) = 1 - e^-λa^ where P(a) is the seroprevalence at age a, A is the force of infection assumed constant over age and e^-λa^ is the probability of remaining seronegative up to age a. The model has fit separately to the data from the in-house ELISA and the Panbio assay using Bayesian inference via Stan in R. Observed age-stratified seroprevalence proportions and binomial sampling distributions were used to model the likelihood. We used weakly informative priors for FOI (λ), and posterior distributions were obtained via Markov Chain Monte Carlo (MCMC) sampling. Posterior medians and 95% credible intervals (CrI) were used to summarize the estimates of FOI and seroprevalence.

## Results

### Age stratified dengue seroprevalence of the children

2160/4161 (51.91%) children of the study were males and 2001 (48.09%) were females. The median age was 10 years (IQR 7 to 13 years). The sero-status of the children was assessed by the in-house DENV-specific IgG ELISA. Accordingly, the overall seropositivity rate of the cohort was 72.34% (3010/4161), with DENV seropositivity rates gradually rising from 56.93% in children aged 4 to 5 years to 82.44% in children aged 14 to 16 years (Table 1). A significant increase in the age stratified seroprevalence rate was seen (Spearman’s r=1.0, p=0.003) (Figure 1A). The annual seroconversion rate was estimated to be 2.3% (±0.27) per annum. There was no difference (p=0.37) in age stratified seroprevalence rates in females compared to males (Figure 1B, Supplementary table 1) in this cohort.

**Figure 1:**
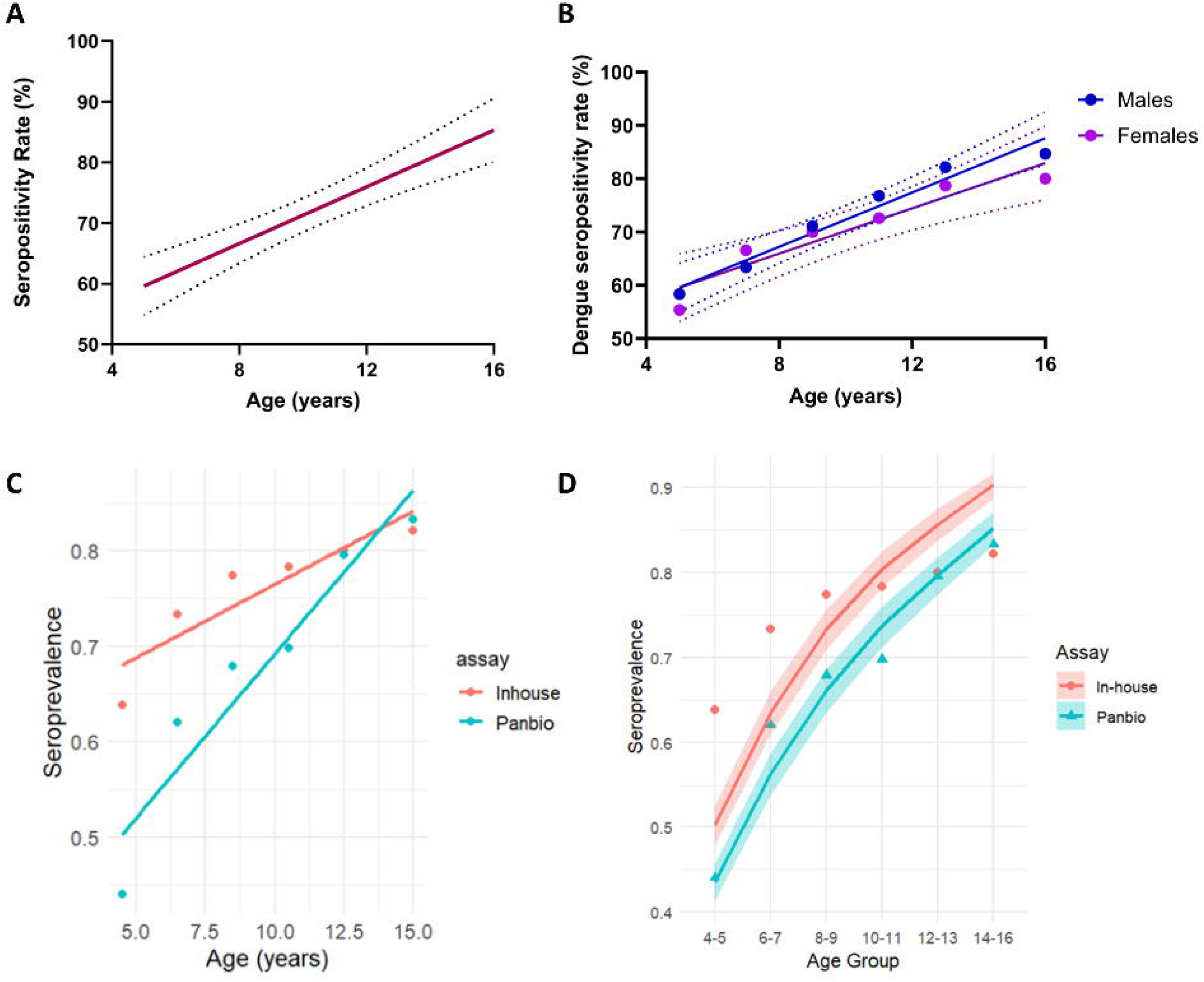
Age stratified seropositivity rates in children. The dengue IgG levels were measured in 2160 males and 2001 females between the ages 4 to 16 years, from the Gampaha district using the in-house DENV specific IgG ELISA, and dengue seropositivity rates correlated with the age (A). The dengue antibody positivity rates were also correlated with age in males (blue) and purple (dark pink) (B). The dengue IgG seroprevalence for the in-house ELISA and the commercial Panbio indirect ELISA. Solid lines represent the best-fit simple linear regression models for each assay, showing the relationship between age and seropositivity. A statistically significant interaction between age and assay type was observed (p = 0.01), indicating differences in the age-seroprevalence slopes between assays (C). The observed and estimated age-stratified seroprevalence using the in-house DENV-specific IgG ELISA and the commercial Panbio assay (D). Dots and triangles represent observed seropositivity proportions by age group, while shaded curves represent Bayesian model-based estimates of seroprevalence with 95% credible intervals. The force of infection (FOI) was estimated using the Bayesian catalytic model.

**Table 1:** Age stratified seroprevalence rates of children in Gampaha district, Sri Lanka. The presence of DENV-specific IgG antibodies was measured in 4161 children aged 4 to 16 years of age, by using an in-house DENV IgG ELISA.

| **Age Group**<br>**(years)** | **Total Number in Each**<br>**Age Group** | **Seropositivity Rate n (%)** | **Seronegativity Rate n (%)** |
| --- | --- | --- | --- |
| 4-5 | 613 | 349 (56.93%) | 264 (43.07%) |
| 6-7 | 643 | 418 (65.01%) | 225 (34.99%) |
| 8-9 | 722 | 509 (70.50%) | 213 (29.50%) |
| 10-11 | 687 | 513 (74.67%) | 174 (25.33%) |
| 12-13 | 647 | 521 (80.53%) | 126 (19.47%) |
| 14-16 | 849 | 700 (82.45%) | 149 (17.55%) |
| **Total** | **4161** | **3010 (72.34%)** | **1151 (27.66%)** |

### Comparison of the in-house DENV-specific IgE assay with the Panbio dengue indirect IgG assay

As the Panbio indirect dengue IgG assay had been widely used in determining dengue seroprevalence in Sri Lanka and elsewhere [9, 18], we compared the age stratified seroprevalence rates given by the in-house IgG assay with that of the Panbio indirect IgG assay in a sub-cohort of children (n=1374). While 1047 (76.2%) children were shown to be dengue seropositive by the in-house DENV-specific IgG ELISA, the Panbio indirect IgG assay gave a positive result for 941/1374 (68.48%) children, with an equivocal result for 14/1374 children. To assess agreement between the two assays (how consistently they classify the same individuals) Cohen’s Kappa statistic used, and it was 0.33 (95% CI 0.27-0.38) suggested fair agreement between the two assays in classifying dengue seropositivity.

To assess if there’s a significant difference in the number of positive results between the two assays, McNemar chi square test showed a significant difference in seropositivity classification between the in-house DENV-specific IgG ELISA and the Panbio Indirect IgG ELISA (χ² = 25.85, p < 0.001), with the in-house assay classifying a greater number of individuals as seropositive. Further, a statistical interaction test using linear regression showed a significant interaction between age and assay type (p = 0.01), indicating that the relationship between age and seroprevalence differs significantly between the Panbio and in-house assays (Figure 1C). The sensitivity of the in-house IgG ELISA was 91.8 % with a specificity of 75% (supplementary methods). In contrast, the sensitivity of the Panbio IgG ELISA was 83.7% with a specificity of 87.5%. Therefore, the in-house IgG ELISA had higher sensitivity than the Panbio ELISA in detecting multitypic infections (100% detection), with better correlation with the FRNT assay.

Age-stratified seroprevalence was modeled using a Bayesian catalytic framework assuming a constant force of infection (FOI). The estimated FOI was 0.16 (95% credible interval [CrI]: 0.14–0.17) for the in-house DENV-specific IgG ELISA and 0.13 (95% CrI: 0.12–0.14) for the commercial Panbio indirect IgG ELISA, indicating a slightly higher estimated rate of DENV exposure when using the in-house assay. Estimated seroprevalence curves closely matched the observed age-stratified seroprevalence proportions for both assays. However, the in-house assay consistently showed higher seroprevalence across all age groups compared to the Panbio assay. The model-derived seroprevalence trajectories and their 95% credible intervals visually confirmed these differences, supporting a higher cumulative detection sensitivity by the in-house assay (Figure 1D).

### Dengue and zika exposure rates in dengue seropositive children

Using a Luminex based bead array, which uses antibodies specific to the EDIII of the four serotypes of the DENV and the EDIII of ZIKV [17], we proceeded to further characterize exposure rates to different DENV serotypes and Zika. These assays were carried out in 20% (n=604), DENV seropositive children, and this sub-cohort was selected randomly to proportionately represent all age groups and gender of children included in the study. The number of children tested in each age group for Luminex assays is shown in Supplementary table 2.

258 (42.7%) of this DENV seropositive sub-cohort were found to have been infected with only one DENV in the past (monotypic dengue infection), while 209 (34.89%) were found to be infected with many DENV serotypes (multi-typic infection). Some children who had past monotypic dengue infection were also found to have been infected with zika and some children who had a past muti-typic dengue infections have also been exposed to zika in the past (Table 2). 20 (3.33%) children, only had antibodies to the ZIKV, indicating that the in-house DENV-IgG ELISA, would detect cross-reactive flavi-virus antibodies. Of the 278 individuals (including those who had a monotypic DENV infection and zika), the main serotype that they had been exposed to was DENV2 (56.83%), followed by DENV1 (30.57%). 37 (6.13%) individuals who gave a positive response with the in-house DENV IgG ELISA, were found to have a negative response with the Luminex assay.

**Table 2:** Exposure to different DENV serotypes and zika in a sub-cohort of children in the Gampaha district. The presence of antibodies to different DENV serotypes and ZIKV was measured in 604 children by using a Luminex assay which uses antigens (EDIII), which can differentiate antibody responses to different DENV serotypes and ZIKV.

| Exposure to past DENV or ZIKV | Number of children who tested positive<br>N (%) | Serotype | Exposure to different DENV serotypes<br>N (%) |
| --- | --- | --- | --- |
| Monotypic | 258 (42.72) | DENV1 | 78 (30.23) |
|  |  | DENV2 | 148 (57.36) |
|  |  | DENV3 | 19 (7.36) |
|  |  | DENV4 | 13 (5.04) |
| Monotypic + ZIKV | 20 (3.31) | DENV1 | 7 (35.00) |
|  |  | DENV2 | 10 (50.00) |
|  |  | DENV3 | 3 (15.00) |
|  |  | DENV4 | 0 |
| Multitypic | 209 (34.60) |  | Cannot be determined |
| Multitypic + ZIKV | 60 (9.93) |  | Cannot be determined |
| ZIKV | 20 (3.31) |  | Not applicable |
| Negative | 37 (6.13) |  | Not applicable |

### Differences in age-stratified exposure to different DENV serotypes and ZIKV

There was no difference in the proportion of children who had a monotypic vs a multi-typic dengue infection (Figure 2A). However, there was a significant increase (Spearman’s=0.62, p=0.026) in monotypic infections with age, suggesting that many seronegative children were continuously exposed to DENV (Fig 2B). We also observed an inverse correlation between exposure to ZIKV and age (Spearmans’s r=-0.72, 0=0.007) (Fig 2B). However, there were very few children included in the age groups 4 to 6 years of age, compared to older age groups. There was no difference in the proportion of children who had experienced multi-typic infections with age.

**Figure 2:**
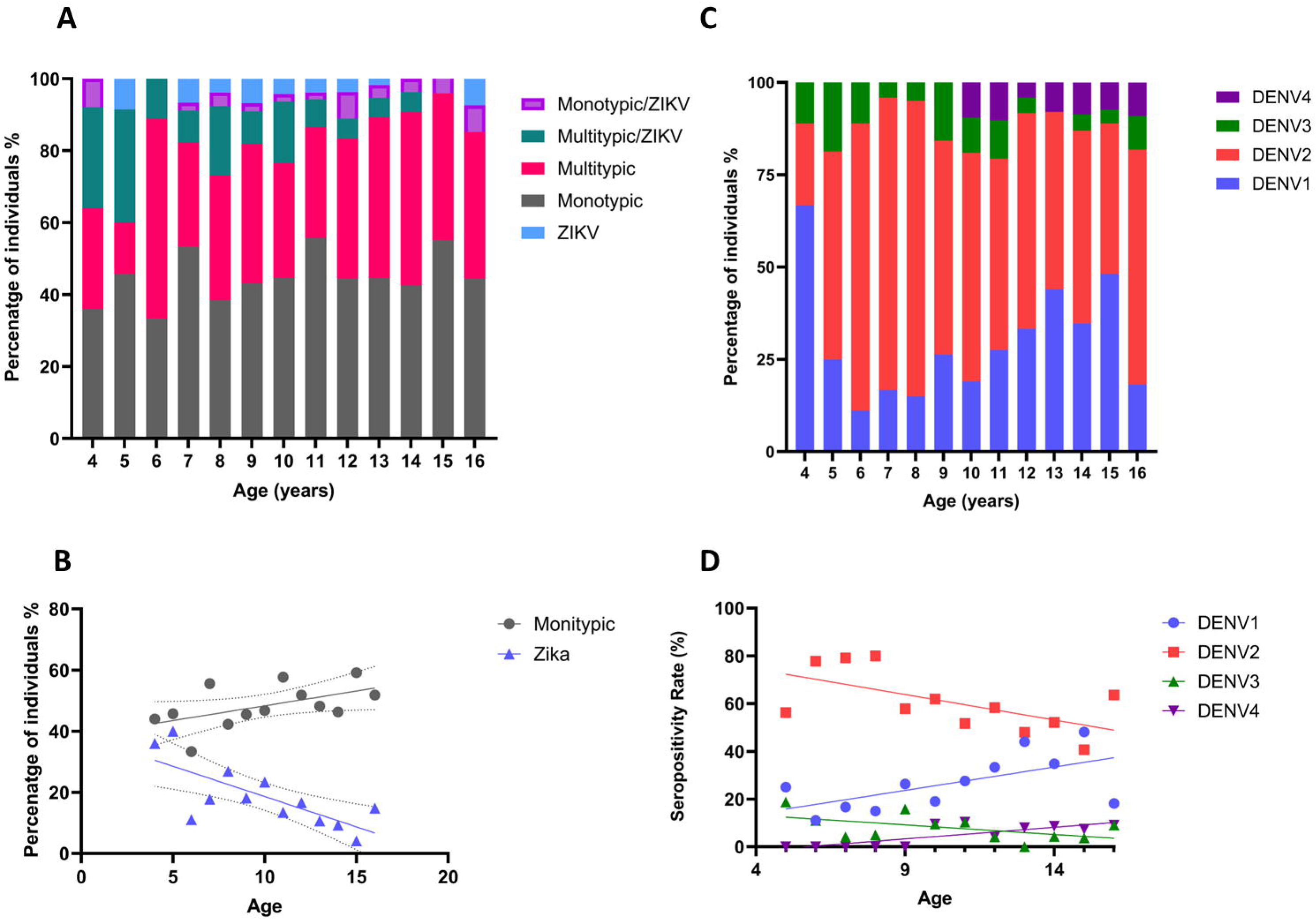
Differences in age-stratified exposure to different DENV serotypes and ZIKV. The proportion of children in different age groups having a past exposure to only one DENV serotype (monotypic infection), or several DENV serotypes (muti-typic infections), ZIKV or a combination of monotypic, multi-typic infection and ZIKV was assessed using a multiplexed, microsphere-based serological assay (Luminex assay) (A). The presence of a monotypic infection (grey) or past exposure to ZIKV (blue), was correlated with the age of the children (B). The multiplexed, microsphere-based serological assay (Luminex assay) was also used to identify the DENV serotype in those with monotypic infection in different age groups (C). The exposure to different DENV infections in children with monotypic infection were correlated with age (D). Spearman rank order correlation coefficient was used to evaluate the correlation between the age, monotypic and ZIKV past infection rates in different age groups, between the infection rates in different DENV serotypes. All tests were two sided.

Of the children with monotypic infections, DENV2 was the most prominent (Figure 2C). DENV4 exposure was not detected in any of the children under the age of 10 years. While there was no significant correlation was seen in exposure to DENV2 and DENV3 with age, both exposure to DENV1 (Spearman’s r=0.63, p=0.03) and DENV4 (Spearman’s r=0.69, p=0.01), significantly increased with age (Fig 2D).

### Relationship between circulating DENV serotypes and infections seen in children

Our laboratory has been carrying out surveillance of DENV serotypes in the Colombo district (district adjacent to the Gampaha district) from 2009 until 2024 and has been published [7, 19, 20]. The data from these surveillance activities, previously published, were used to determine the changes in DENV serotypes throughout this period (Figure 3). Our data shows that DENV1 was the predominant serotype from 2009 to 2016, DENV2 from 2016 to 2019, with DENV3 emerging toward the end of 2019 (Figure 3). During the year 2020 and 2021, the incidence of dengue markedly reduced, and only limited DENV surveillance activities were carried out. In 2021 although DENV2 was still the predominant serotype, it was shown to be gradually replaced by DENV3 since 2023 [19] (Figure 3). Since DENV2 was the predominant serotype from 2016 to 2023 the main serotype causing past infection in children with monotypic dengue infections were seen to be DENV2. DENV1 was seen in 30.57% of children with monotypic infections, again, as it had been circulating from 2009 and again in recent years. DENV4 had been detected in Colombo until Q2 of 2018 detected in 2 to 8% of individuals with dengue infections. It was again detected in Q4 of 2019 in 2.6% of individuals. In our cohort, no one under the age of 10 with monotypic DENV infection was seen to be infected with DENV4 in the past.

**Figure 3:**
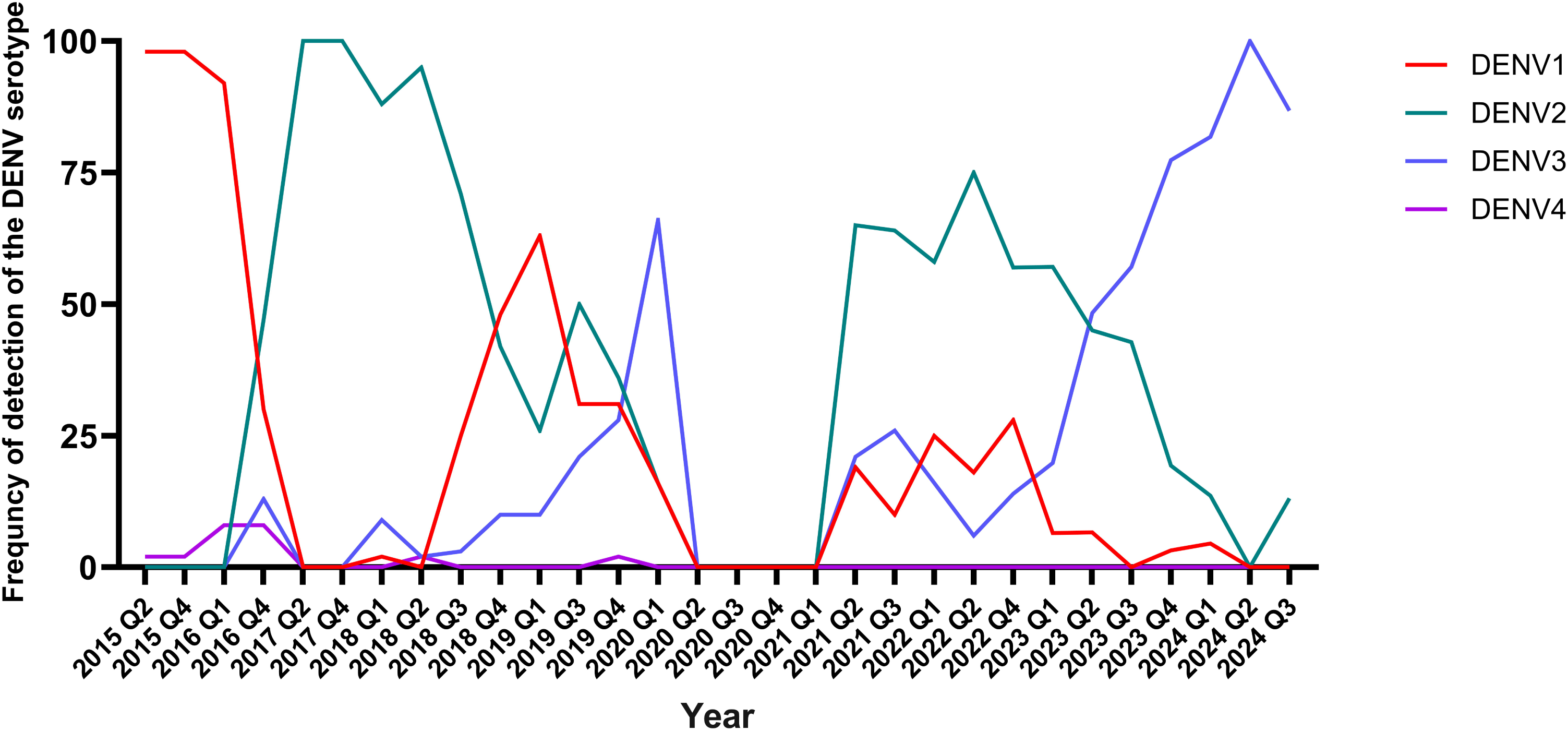
Changes in the circulating DENV serotypes in Colombo, Sri Lanka from 2015 to January 2024.

## Discussion

In this study we have determined the age stratified dengue seroprevalence in a large cohort of children in the Gampaha district, which reports the highest number of dengue infections in Sri Lanka, along with the Colombo district [6]. Our overall seroprevalence was 72.34% which was comparable urban populations of Malaysia, Dhakka in Bangladesh, São Paulo (Brazil) and Cambodia, but lower than in Thailand, Laos, the Philippines and [10, 21–23]. The estimated FOI was 0.16 for Gampaha district in this study, which was higher than previously reported by us, in Sri Lanka in year 2022, where DENV seroprevalence was assessed using the Panbio ELISA [9, 10]. The Panbio ELISA has shown to be less sensitive in detecting previous exposure to the DENV, especially in children with monotypic dengue infection, as shown in Cebu, Philippines [24]. In this study, as our in-house IgG ELISA was found to be more sensitive than the Panbio IgG ELISA, and we possibly got higher seroprevalence rates than previously reported. However, the areas chosen for this study, were the areas which reported the highest dengue incidence in recent years, as assessing the age stratified seroprevalence in these areas, was carried out a part of a clinical trial to evaluate the effectiveness of a spatial repellent [6]. Therefore, unlike our previous study for assessing age-stratified DENV seroprevalence [9], which included urban, rural and estate areas, as areas reporting the highest incidence was chosen for this study, it might overestimate the FOI in the whole district.

Females have shown to have a higher risk of developing severe dengue, and since higher case fatality rates have been reported in females [25–27], we proceeded to determine if there were differences in age stratified seroprevalence rates in females compared to males. Age-stratified seroprevalence rates were similar in females and males despite the observation of higher severe dengue incidence and mortality reported among females. The reasons for higher disease severity and mortality in females does not appear to be associated with higher exposure in this group, but likely due to yet to be defined risk factors or gender-based case management differences.

Although we and others had conducted several studies to determine age-stratified seroprevalence rates in different districts in Sri Lanka and over time in Colombo [7–9], this is the first time we estimated exposure rates to different DENV serotypes; we were able to define serotypes in primary infections represented by a monotypic response, whereas individuals with a multi-typic response probably represented primarily secondary infections with some primary infections with cross reactive antibodies. This cohort will be sampled annually to estimate incidence rates to evaluate the protective efficacy of a spatial repellent based vector control intervention, in cohort members who naïve or monotypic because of the difficulty associated with identifying new secondary infections serologically. 42.7% of dengue seropositive children were found to have experienced only one DENV infection in the past, which again indicated higher transmission rates than Northern, Western and Eastern regions of India, but similar to the trends seen in countries such as Indonesia [28, 29]. Although the seropositivity rates in the children in our study (72.34%), was similar to seropositivity rates seen in Southern India in children of the same age (69.6%), most children had a multi-typic infection in Southern India, indicating higher transmission rates than in our cohort. This highlights the importance of characterizing the proportion of past dengue infections, which are monotypic or multi-typic in nature, as it gives more granularity regarding dengue transmission dynamics.

Due to the highly cross-reactive nature of flavivirus antibodies, the presence of IgG antibodies to the DENV [13, 30], could also indicate infection with cross-reactive flaviviruses such as ZIKV, WNV and JEV which has been shown to circulate in Sri Lanka [31–33]. Furthermore, seropositivity rates alone do not indicate the exposure rates to the DENV, as it would not indicate if an individual has been infected once or several times. In this study, we were also able to differentiate between monotypic and multi-typic infections, we were also able to identify the past infecting serotype in children with monotypic infections. DENV2 (56.83%) was the main DENV serotype that had infected most children followed by DENV1 (30.57%). These findings are consistent with our surveillance data carried out over the years in the Colombo district (adjacent district), which showed that DENV-2 was indeed the predominant circulating serotype from 2016 until end of 2022 [20] (Figure 3). There was a trend toward a reduction of exposure to DENV-2 with age, which could be due to individuals being infected with other DENV2 resulting in multi-typic infections.

DENV-4 was not identified in our surveillance after 2018, with it been detected in very low frequency in the 4^th^ quarter of 2019. In consistent with our surveillance data, we did not identify children under the age of 10, who had been infected with DENV4. DENV3 emerged in 2023, after it was not detected since 2008 (although it was detected for a brief period during the 4^th^ quarter of 2019) [19]. We saw an equal proportion of children exposed to DENV3 in all ages. Therefore, in the absence of past surveillance data, determining exposure rates to different DENV serotypes in different age groups by these multiplexed, microsphere-based serological assays (Luminex assays), would enable us to characterize the past exposure history and to get an idea regarding the transmission of DENVs in a particular geographic location in relation to time.

Although we and others had studied seroprevalence of dengue over many years and in many geographical locations, the seropositivity rates for ZIKV had not been characterized. We found that 16.5% of children had also evidence of past infection with ZIKV with 3.33% of children only reporting infection with ZIKV, despite the in-house DENV-specific IgG giving a positive result. As virological surveillance for ZIKV has not been carried out in Sri Lanka previously, there is no data regarding when ZIKV was first introduced to Sri Lanka or how long the virus has been circulating here. However, the first time the ZIKV was identified in Sri Lanka was in patients presenting with a dengue-like illness during 2017 to 2018 to a tertiary care hospital in Sri Lanka [31], which does not obviously indicate that ZIKV was not circulating prior to 2017. In this study, ZIKV was detected in 1% (6/595) samples by real-time PCR [31]. Interestingly, we saw that infection rates with ZIKV significantly declined with age. Although the reasons for this are not clear, it is possible that prior multi-typic infection with DENV reduces the risk of infection with the ZIKV. In fact, it has been shown that pre-existing, higher antibody titters to the DENV was associated with a lower risk of symptomatic Zika [34]. As ZIKV infection was only seen in younger children with lower infection rates with ZIKV in older children, it is likely that ZIKV was introduced to Sri Lanka recently, as older children who were dengue seropositive, possibly with multi-typic infections were protected from ZIKV. Therefore, as the rate of DENV infection increases with age, this could lead to the risk of infection with ZIKV, which would explain our observations.

In conclusion, we found that 72.3% of children were seropositive for dengue, in Gampaha district in Sri Lanka, which is one of the districts that report the highest number of cases. The dengue seropositivity rate significantly increased with age, although 42.7% of children having been infected with only one DENV in the past (monotypic infection). The predominant infecting serotype was DENV2 followed by DENV1, which is consistent with the DENV serotype circulating data in the last 10 years. Interestingly, infection rates with ZIKV significantly declined with age, suggesting that prior immunity to DENV may reduce the risk of ZIKV infection. This data shows that in addition to age-stratified seroprevalence data, understanding the proportion of monotypic and multi-typic infections and exposure to other flaviviruses, is likely to inform better decision making in implementing vaccines and novel vector control strategies.

## Funding

This study was funded by Unitaid (2018-29-UND). Unitaid is a global health organization that saves lives by making new health products available and affordable for people in low- and middle-income countries. Unitaid works with partners to identify innovative treatments, tests and tools, help tackle the market barriers that are holding them back and get them to the people who need them most – fast. Since Unitaid was created in 2006, the organization has unlocked access to more than 100 groundbreaking health products to help address the world’s biggest health challenges, including HIV, TB and malaria; women’s and children’s health; and pandemic prevention, preparedness and response. Every year, more than 300 million people benefit from the products Unitaid has helped roll out. Unitaid is hosted by the World Health Organization.

## Data Availability

Data is available in the manuscript, figures and the supporting files.

## Notes

### Competing Interest Statement

GNM has served in advisory capacities on dengue for Johnson & Johnson, Novartis, Takeda and Abbott.

