## Supplementary methods for "Dissecting age-stratified immunity to different dengue virus serotypes and Zika viruses among children in a highly endemic region in Sri Lanka"

**Development and validation of an in-house IgG assay for detection of dengue virus (DENV) specific IgG antibodies for identification of individuals with a past dengue infection**

**Samples used for the development**

Serum samples from 76 individuals were used for the development of this assay. Foci reduction neutralization assays (FRNTs) were previously carried out on these samples to determine the dengue serostatus [1]. Out of the 76 samples, 17 had dengue monotypic profiles, 35 had multitypic profiles and 24 were seronegative for dengue.

**Propagation of viruses used for Coating**

DENV1 WestPac74, DENV2 S16803, DENV3 CH53489 and DENV4 TVP-376 (donated by Prof. Aravinda de Silva) were propagated in a Vero-81 cell line. The harvested viruses were then subjected to foci forming assays (FFAs) to determine the concentrations in foci forming units/ml (ffu/ml). These viruses were then used for the subsequent in-house dengue IgG ELISA assays that we carried out.

**Development and Optimization of the Dengue IgG ELISA**

The in-house ELISA was developed by coating the plates with viruses from the four DENV serotypes (DENV1, DENV2, DENV3, and DENV4) and the DENV specific IgG in patient serum was captured. Coating concentrations of the DENV viruses, blocking concentration, serum dilutions, secondary antibody concentration and incubation times were optimized using checkerboard titrations.

The ELISA was carried out on 96-well high binding ELISA plates (PierceTM, Cat: 15031) which was first coated with the four DENV viruses, that were diluted in PBS at a coating concentration of 250 ffu/ml each. The plates were incubated overnight at 4°C. On the following day, plates were washed once with 250µl/well of PBS and then the wells were blocked with 250µl/well of 2% Bovine Serum Albumin (BSA) in PBS (blocker) for 1.5 hours at room temperature. Serum samples were diluted at 1:250 in blocker, and the diluted serum was added in duplicates into the respective wells at 100µl/well after which the plate was incubated for 1 hour at room temperature. The wells were then washed thrice with wash buffer (PBS + 0.05% Tween 20). Then, 100µl of goat anti-human IgG biotinylated antibody (Mabtech, Sweden, Cat: 3820-4-250) (diluted at 1:1000 in PBS containing 2% BSA) was added into each well and the plate was incubated for an hour at room temperature. After incubation, the plate was washed thrice and 100µl of diluted Streptavidin-HRP (Mabtech, Sweden, Cat: 3310-9) (1:1000 in PBS containing 2% BSA) was added to each well and incubated for 30 minutes at room temperature in the dark, followed by five washes. The colour development was carried out by adding 80 µl of TMB ELISA substrate solution (Mabtech, Sweden, Cat: 3652-F10) to each well and incubating for 30 minutes in the dark. The reaction was stopped with 80 µl of 1N HCl (stop solution) and the absorbance was read at 450nm using the MPSCREEN MR-96A ELISA reader. Blocker was added to duplicate wells as blank for the assay. This ELISA was carried out on the 4161 samples to determine their dengue serostatus.

**Establishment of cut-off to determine positives in the in-house IgG ELISA**

Stastistical Analysis was carried out using GraphPad Prism version 10.1 Non-parametric tests were carried out, since the data was not normally distributed. A receiver-operating characteristic (ROC) curve was generated for the developed in-house IgG ELISA to determine the threshold value that distinguishes positive from negative results and also considering the sensitivity and specificity percentages of the assay.

Results of the positive samples (n=52) determined by the FRNTs were used to determine the cut-off. ROC analysis was conducted to evaluate the accuracy of the assay and identify the cut-off value that gives the highest sensitivity and specificity. The ROC curve is shown in Figure 1, and the area under the curve was 0.8686 ± 0.050 with a 95% confidence interval of 0.7692 to 0.9680. The cut-off selected for determining positivity was an OD of 0.5950 with a sensitivity of 92.3% and a specificity of 75%. This cut-off showed that 48/52 FRNT positive samples were positive using the IgG ELISA, where 4/17 monotypics were negative, and out of the 24 FRNT negative samples 18 were negative by the ELISA and 6 were positive. The cut-off of 0.5950 was used for the tests that followed.

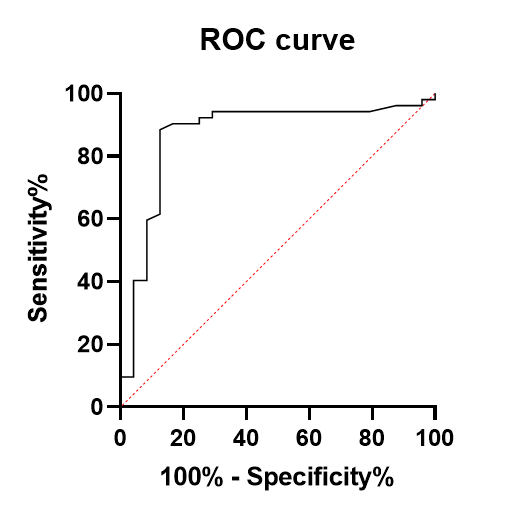

Figure 1: Receiver operating characteristic (ROC) analysis was carried out for the developed ELISA using positive samples confirmed by the foci reduction neutralization test (FRNT). The graph demonstrates the ROC curve.

**Validation of the results of the in-house DENV-specific IgG ELISA with FRNT and Panbio IgG ELISA assays**

In order to determine the accuracy of the in-house DENV-specific IgG ELISA, we compared the positive results given by this assay with the results of the FRNT and the PanBio IgG ELISA. The Panbio ELISA was carried out according to the manufacturer’s instructions, where those that had Panbio Units <9 were classified as negative, Panbio Units 9-11 were classified as equivocal and those with Panbio Units >11 were classified as being positive for a past dengue infection.

Of the 33 samples which gave a multi-typic response with the FRNT assay, all 33 gave a positive response with the in-house IgG ELISA, while 2 samples gave a negative result with the Panbio IgG ELISA (supplementary table 1). Of the 16 monotypic infections identified with the FRNT assay, the in-house IgG assay gave a positive response for 12/16 (it was 10/16 with the Panbio assay). Of the 24 negative samples by the FRNT assay, the IgG assay gave a positive response to 6, while the Panbio ELISA gave a positive response to 3.

| FRNT | | Panbio | | | | IgG | |
| --- | --- | --- | --- | --- | --- | --- | --- |
| n=73 | | **Positive** | **Equivocal** | **Negative** | **Positive** | | **Negative** |
| Multitypic | 33 | 31/33 | - | 2/33 | | 33/33 | - |
| Monotypic | 16 | 10/16 | - | 6/16 | | 12/16 | 4/16 |
| Negative | 24 | 3/24 | - | 21/24 | | 6/24 | 18/24 |

**Supplementary table 1: Comparison of the results using the PanBio Indirect IgG ELISA and the in-house IgG ELISA using samples with known dengue serostatus confirmed by FRNT.**
