## Supplementary data for "Dissecting age-stratified immunity to different dengue virus serotypes and Zika viruses among children in a highly endemic region in Sri Lanka"

**Supplementary results**

| **Age Group** | **Total Number in Each Age Group** | **Males** | | | **Females** | | |
| --- | --- | --- | --- | --- | --- | --- | --- |
|  |  | **Number in Each Age Group** | **Seropositivity Rate**  **N (%)** | **Sero-negativity Rate**  **N (%)** | **Number in Each Age Group** | **Seropositivity Rate**  **N (%)** | **Sero-negativity Rate**  **N (%)** |
| 4-5 | 613 | 324 | 189 (58.33%) | 135(41.67%) | 289 | 160 (55.36%) | 129 (44.63%) |
| 6-7 | 643 | 341 | 217 (63.36%) | 124 (36.36%) | 302 | 201 (66.56%) | 101 (33.44%) |
| 8-9 | 722 | 369 | 262 (71.10%) | 107 (29.00%) | 353 | 247 (70.00%) | 106 (30.55%) |
| 10-11 | 687 | 340 | 261 (76.77%) | 79 (23.23%) | 347 | 252 (72.62%) | 95 (27.38%) |
| 12-13 | 647 | 342 | 281 (82.16%) | 61 (17.84%) | 305 | 240 (78.69%) | 65 (21.31%) |
| 14-16 | 849 | 444 | 376 (84.68%) | 68 (18.10%) | 405 | 324 (80.00%) | 81 (20.00%) |
| **Total** | **4161** | **2160** | **1586 (73.43%)** | **574 (26.57%)** | **2001** | **1424 (71.17%)** | **577 (28.84%)** |

Supplementary table 1:

| Age | Antigen | Number | Total | Percentages |
| --- | --- | --- | --- | --- |
| 4 | Monotypic | 9 |  | 36.00 |
|  | Multitypic | 7 |  | 28.00 |
|  | Multitypic/ZIKV | 7 |  | 28.00 |
|  | Monotypic/ZIKV | 2 |  | 8.00 |
|  | ZIKV | 0 |  | 0.00 |
|  | Negative | 6 | 31 | 24.00 |
| 5 | Monotypic | 16 |  | 45.71 |
|  | Multitypic | 5 |  | 14.29 |
|  | Multitypic/ZIKV | 11 |  | 31.43 |
|  | Monotypic/ZIKV | 0 |  | 0.00 |
|  | ZIKV | 3 |  | 8.57 |
|  | Negative | 6 | 41 | 17.14 |
| 6 | Monotypic | 9 |  | 33.33 |
|  | Multitypic | 15 |  | 55.56 |
|  | Multitypic/ZIKV | 3 |  | 11.11 |
|  | Monotypic/ZIKV | 0 |  | 0.00 |
|  | ZIKV | 0 |  | 0.00 |
|  | Negative | 5 | 32 |  |
| 7 | Monotypic | 24 |  | 53.33 |
|  | Multitypic | 13 |  | 28.89 |
|  | Multitypic/ZIKV | 4 |  | 8.89 |
|  | Monotypic/ZIKV | 1 |  | 2.22 |
|  | ZIKV | 3 |  | 6.67 |
|  | Negative | 5 | 50 | 11.11 |
| 8 | Monotypic | 20 |  | 38.46 |
|  | Multitypic | 18 |  | 34.62 |
|  | Multitypic/ZIKV | 10 |  | 19.23 |
|  | Monotypic/ZIKV | 2 |  | 3.85 |
|  | ZIKV | 2 |  | 3.85 |
|  | Negative | 2 | 54 | 3.85 |
| 9 | Monotypic | 19 |  | 43.18 |
|  | Multitypic | 17 |  | 38.64 |
|  | Multitypic/ZIKV | 4 |  | 9.09 |
|  | Monotypic/ZIKV | 1 |  | 2.27 |
|  | ZIKV | 3 |  | 6.82 |
|  | Negative |  | 44 | 0.00 |
| 10 | Monotypic | 21 |  | 44.68 |
|  | Multitypic | 15 |  | 31.91 |
|  | Multitypic/ZIKV | 8 |  | 17.02 |
|  | Monotypic/ZIKV | 1 |  | 2.13 |
|  | ZIKV | 2 |  | 4.26 |
|  | Negative | 4 | 51 | 8.51 |
| 11 | Monotypic | 29 |  | 55.77 |
|  | Multitypic | 16 |  | 30.77 |
|  | Multitypic/ZIKV | 4 |  | 7.69 |
|  | Monotypic/ZIKV | 1 |  | 1.92 |
|  | ZIKV | 2 |  | 3.85 |
|  | Negative | 4 | 56 | 7.69 |
| 12 | Monotypic | 24 |  | 44.44 |
|  | Multitypic | 21 |  | 38.89 |
|  | Multitypic/ZIKV | 3 |  | 5.56 |
|  | Monotypic/ZIKV | 4 |  | 7.41 |
|  | ZIKV | 2 |  | 3.70 |
|  | Negative | 1 | 55 | 1.85 |
| 13 | Monotypic | 25 |  | 44.64 |
|  | Multitypic | 25 |  | 44.64 |
|  | Multitypic/ZIKV | 3 |  | 5.36 |
|  | Monotypic/ZIKV | 2 |  | 3.57 |
|  | ZIKV | 1 |  | 1.79 |
|  | Negative | 1 | 57 | 1.79 |
| 14 | Monotypic | 23 |  | 42.59 |
|  | Multitypic | 26 |  | 48.15 |
|  | Multitypic/ZIKV | 3 |  | 5.56 |
|  | Monotypic/ZIKV | 2 |  | 3.70 |
|  | ZIKV | 0 |  | 0.00 |
|  | Negative | 1 | 55 | 1.85 |
| 15 | Monotypic | 27 |  | 55.10 |
|  | Multitypic | 20 |  | 40.82 |
|  | Multitypic/ZIKV | 0 |  | 0.00 |
|  | Monotypic/ZIKV | 2 |  | 4.08 |
|  | ZIKV | 0 |  | 0.00 |
|  | Negative | 2 | 51 | 4.08 |
| 16 | Monotypic | 12 |  | 40.74 |
|  | Multitypic | 11 |  | 40.74 |
|  | Multitypic/ZIKV | 0 |  | 0.00 |
|  | Monotypic/ZIKV | 2 |  | 7.41 |
|  | ZIKV | 2 |  | 7.41 |
|  | Negative | 0 | 27 | 0.00 |

Supplementary table 2:
